# Anti-Gephyrin antibodies: A novel specificity in patients with systemic sclerosis and lower bowel dysfunction

**DOI:** 10.1101/2022.08.26.22279277

**Authors:** ZH McMahan, S Kulkarni, F Andrade, J Perin, C Zhang, JE Hooper, FM Wigley, A Rosen, PJ Pasricha, L Casciola-Rosen

## Abstract

**Objective:** Autoantibodies are clinically useful in phenotyping patients with systemic sclerosis (SSc). GI function is regulated by the enteric nervous system (ENS) and commonly impaired in SSc suggesting that the SSc autoimmune response may target ENS antigens. We sought to identify novel anti-ENS autoantibodies with an aim to clinically phenotype SSc GI dysfunction.

**Methods:** Serum from a SSc patient with GI dysfunction but without defined SSc-associated autoantibodies was used for autoantibody discovery. Immunoprecipitations performed with murine myenteric plexus lysates were on-bead digested and autoantigens were identified by mass spectrometry. Prevalence and clinical features associated with novel autoantibodies were evaluated in a SSc cohort using regression analyses. Expression of gephyrin in human GI tract tissue was examined by immunohistochemistry.

**Results:** We identified gephyrin as a novel SSc autoantigen. Anti-gephyrin antibodies were present in 9% of SSc patients (16/188) and absent in healthy controls (0/46). Anti-gephyrin antibody-positive patients had higher constipation scores [1.00 vs. 0.50;p=0.02] and were more likely to have severe constipation and severe distention/bloating [46% vs. 15%;p=0.005; 54% vs. 25%;p=0.023, respectively]. Anti-gephyrin antibody levels were significantly higher among patients with severe constipation [0.04 vs. 0.00;p=0.001] and severe distention and bloating [0.03 vs. 0.004;p=0.010]. Severe constipation was associated with anti-gephyrin antibodies even in the adjusted model. Importantly, gephyrin was expressed in the ENS, which regulates gut motility.

**Conclusion:** Gephyrin is a novel ENS autoantigen that is expressed in human myenteric ganglia. Anti-gephyrin autoantibodies are associated with the presence and severity of constipation in SSc patients.

## INTRODUCTION

Systemic sclerosis (SSc) is an autoimmune rheumatic disease associated with fibrosis, vasculopathy, and internal organ dysfunction. While these common findings unite most SSc patients, clinically distinct profiles exist within SSc that associate tightly with autoantibody status [e.g. anti-RNA polymerase-3 antibodies associate with renal crisis] (1). Given that the well-defined SSc autoantigens are ubiquitously expressed, the close association between clinical phenotypes and specific SSc-autoantibodies raises intriguing questions as to why distinct tissues may be affected among patients with different autoantibodies.

The gastrointestinal (GI) tract is the most commonly involved internal organ in SSc. Patients present with various complications, from mild heartburn to recurrent lower gut pseudo-obstruction. Normal intestinal motility is controlled by neurons and glial cells of the enteric nervous system (ENS) in association with various other cell types in the gastrointestinal tract (2). Thus, correlation between SSc autoantibodies and gut dysmotility would suggest that SSc autoantibodies target autoantigens expressed by the ENS and associated gastrointestinal cells. While autoantibodies are useful in the clinical risk stratification of many SSc complications, their use in GI risk stratification has unfortunately been limited. Functional autoantibodies to the muscarinic-3-receptors (M3R) expressed in the ENS and GI smooth muscle, are reported SSc patients with rapidly progressive lower bowel disease and pseudo-obstruction, but this represents only a small subset of SSc GI patients (<10%) (3). Others showed that sera from SSc patients with significant GI dysfunction have autoantibodies that target diverse proteins within the soma of enteric neurons, though the specific antigens were not identified (5). Finally, Ahmed et al. (6) utilized a cohort of >500 SSc patients to demonstrate that SSc GI burden may correlate with some autoantibody subtypes. In particular, antibodies to centromere and RNA polymerase-3 were associated with a higher SSc GI symptom burden, although in this study the specific GI complications were not defined (6). Interestingly, patients without extractable nuclear antigen antibodies (ENA negative) had a higher SSc GI symptom burden, suggesting that antibodies which are yet to be discovered may associate with SSc GI severity.

Here we identify gephyrin as a novel autoantigen targeted in SSc, and show that gephyrin is expressed by human and murine myenteric neurons. Antibodies against this protein associate with a distinct GI phenotype. Lower GI symptoms, specifically constipation, distention, and bloating are more severe among patients with anti-gephyrin antibodies, and anti-gephyrin antibody levels are significantly higher among SSc patients with more severe constipation.

## METHODS

### Isolation and culture of the longitudinal muscle containing myenteric plexus (LM-MP) from adult murine small intestine (ileum)

LM-MP tissue extracts were prepared as described (Supplemental Methods).

### Identification of antibodies in serum FW-2340

This was performed by immunoprecipitation (IP)/on-bead digest and mass spectrometry (see Supplemental Methods).

### Assays to validate antibodies

Anti-RUVBL1/2 antibodies were confirmed by IP (see Supplemental Methods). Anti-gephyrin antibodies were validated using 3 different assays: (i) IP/blots - these were performed as described (7) using mouse brain lysate as the antigen source (robust levels of gephyrin are expressed in brain). IPs were performed with serum from FW-2340 or a healthy control, followed by blotting with an anti-gephyrin monoclonal antibody (R&D Systems,1:800); (ii) blots performed using purified recombinant human gephyrin (Origene, #TP305986) and (iii) ELISA (see below).

### Anti-gephyrin antibody ELISA

Duplicate ELISA plate wells were coated with 50ng of recombinant human gephyrin (Origene, #TP305986). For each serum, an adjacent well was incubated with PBS (to determine sample background). Wells were washed with PBS/0.05% Tween (PBST), then blocked with 5% BSA/PBST. Primary antibody incubations were performed using sera diluted 1:150 in 1% BSA/PBST (1 hr, RT), followed by incubation with horseradish peroxidase (HRP)-labeled anti-human IgG (Jackson ImmunoResearch, 1:10,000). Color was developed with SureBlue peroxidase reagent (KPL). Reactions were terminated with HCl, and absorbances were read at 450nM. The same anti-gephyrin-positive serum was included as a reference in every ELISA and all absorbances were calibrated relative to it. The cutoff for antibody positivity was determined by assaying sera from 40 healthy controls. The mean + 4SD of these values (0.206 calibrated OD units) was used as the cutoff.

### Immunofluorescence staining of murine LM-MP tissue

Ileal LM-MP tissues from adult male Wnt1-cre:tdTomato lineage fate mapping mice, which we have previously used to label all neural crest-derivatives in the GI tract with the red fluorescent reporter tdTomato, were isolated as described (8). LM-MP tissues were fixed with 4% paraformaldehyde within 30 minutes of isolation, sectioned and immunostained with serum FW-2340 (1:1,000) and a commercial anti-gephyrin antibody (Proteintech, 1:250). Visualization was performed using anti-human 647 (cyan) and anti-rabbit 488 (green) antibodies (both 1:500). Nuclei were stained with DAPI and imaging was performed using confocal microscopy. Tissues were imaged under a 40X oil immersion objective lens, with laser settings selected to ensure no overlap between fluorophores. Images were analyzed using the Fiji ImageJ software.

### Immunohistochemical staining of human GI tract paraffin sections

Human autopsy specimen use was approved by the Johns Hopkins Institutional Review Board (IRB). These were collected through the Johns Hopkins Legacy Gift Rapid Autopsy program (expedites autopsies to preserve tissue integrity). Immunostaining protocols are described (Supplemental Methods and (9)). Matching GI autopsy tissue from patients with neither SSc nor GI disease were used as controls.

## Patient Cohorts

### SSc All-Comers Cohort

This consisted of all SSc patients regardless of GI disease status recruited consecutively during routine clinical visits from April 2016 to August 2017 (N=188). The Johns Hopkins Scleroderma Center Research Registry contains demographic and detailed clinical data from patients at their first clinical encounter and every 6 months thereafter (Supplemental Material). Evidence of patient-reported GI symptoms was determined by the maximum UCLA SCTC GIT 2.0 survey score from the time closest to the serum sample (17).

### Healthy Controls

Serum was obtained from 40 healthy controls.

The Johns Hopkins IRB approved all human subjects’ protocols used, and all individuals provided informed consent.

#### Statistical analysis

Clinical and demographic features were compared between anti-gephyrin antibody positive and negative patients using Chi-square or Fischer’s exact tests, Student’s t-tests or Wilcoxon-Mann Whitney according to the type and distribution of the data. ANOVA was used to compare differences in means across >2 groups. To determine whether anti-gephyrin antibodies associated with disease severity, regression models were used. Adjusted multivariable models were developed, including disease duration and statistically significant variables from the univariate analyses. STATA 15 (STATA Corporation, College Station, Texas) was utilized for all analyses. A p-value of <0.05 was considered statistically significant.

## RESULTS

### Anti-gephyrin antibodies are a novel specificity in patients with SSc and GI disease

To identify ENS autoantigens, we utilized lysate from adult murine LM-MP tissue as the antigen source. Serum from SSc patient FW-2340 was carefully selected to screen for novel autoantibodies based on the following criteria: this patient had severely delayed lower bowel transit (by whole gut scintigraphy), a GI Medsger severity score of 4 (i.e., dependent on total parenteral nutrition), and lacked SSc autoantibodies (per Euroimmun SSc panel). IPs performed with this serum using LM-MP lysates were subjected to on-bead digestion followed by mass spectrometry. This identified two putative autoantibody specificities: RUVBL1/2 (47% coverage) and gephyrin (7% coverage). Antibodies were validated by IVTT-IP (RUVBL1/2), or IP/blotting, blotting and ELISA (gephyrin) as described (Methods and **Figure 1**). As anti-RUVBL1/2 antibodies have been previously reported in SSc and are not associated with SSc GI disease (10), we did not study this specificity further. We instead focused on the novel finding of anti-gephyrin antibodies. While these antibodies were previously reported in one case of Stiff Skin Syndrome (SSS) (11), they have never been described in SSc.

**Figure 1.**
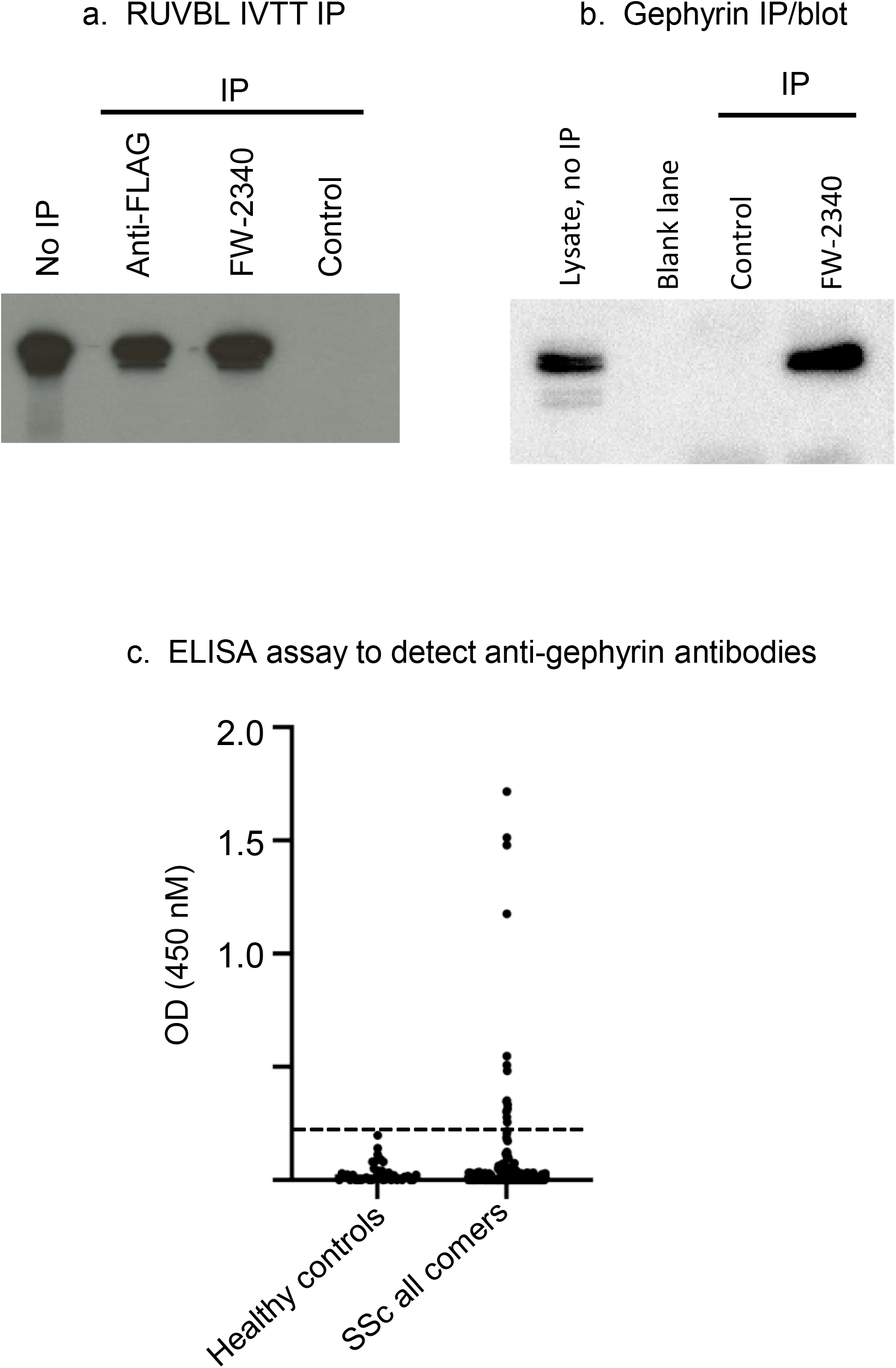
Assays to detect antibodies against RUVBL1/2 and gephyrin. (a): Antibodies against RUVBL1/2 were assayed by immunoprecipitation (IP) using FLAG-tagged 35S-methionine– labeled human RUVBL1/2 as input (see Supplemental Methods). A positive reference IP was performed using an anti-FLAG mouse monoclonal antibody (Sigma). The “Control” lane shows an IP performed using serum from a healthy control individual. FW-2340 is the prototype serum in which anti-gephyrin antibodies were confirmed using this assay. The “No IP” lane shows the input material used for the IPs. (b): Anti-gephyrin antibodies were confirmed in serum FW-2340 using an IP/blot assay, as described in Methods. Lanes marked “Control” and “FW-2340” are as described in (a). No material was loaded in the lane denoted “blank”. (c): Anti-gephyrin antibodies were assayed by ELISA in sera from healthy controls (n=40) and scleroderma all comers (n=188) as described in the Methods section. The dotted line represents the cutoff for assigning a positive anti-gephyrin antibody status (see methods).

### Anti-gephyrin antibodies are prevalent in SSc but not in controls

To determine the prevalence of anti-gephyrin autoantibodies in SSc, we screened a cohort of 188 consecutively recruited patients (**Supplemental Table 1**). We found 16/188 (8.5%) had anti-gephyrin antibodies. Of these, 4/16 (25%) had anti-gephyrin antibodies in the absence of other defined SSc antibodies. None of the healthy controls (0/40) tested positive for anti-gephyrin antibodies (Suppl Fig1C).

### Anti-gephyrin antibodies associate significantly with lower bowel dysfunction

To define the clinical phenotype associated with anti-gephyrin antibodies, we compared antibody-positive and negative SSc patients (**Table 1**). Both groups were similar in terms of age, sex, disease duration, and race. When comparing GI symptom severity, patients with anti-gephyrin antibodies had significantly more constipation, with median scores of 1.00 (0.25, 1.75) compared to 0.50 (0.00, 0.75) in the antibody negative group (p=0.02). Furthermore, anti-gephyrin positive patients were more likely to have severe constipation and severe distention/bloating [46% vs. 15%; p=0.005; 54% vs. 25%; p=0.023, respectively] on the UCLA GIT 2.0.

**Table 1.**
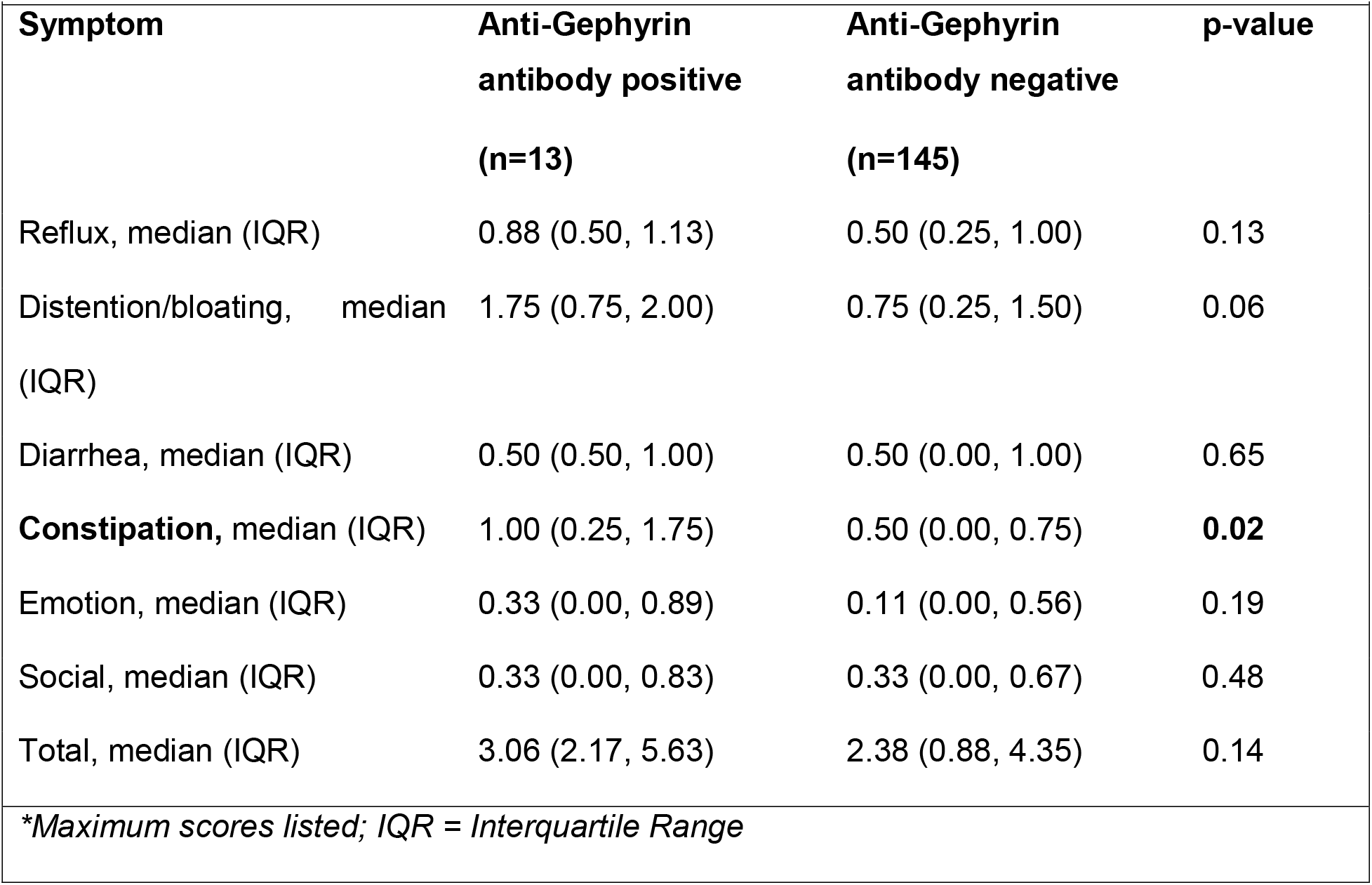
Association between Anti-Gephyrin antibodies and UCLA GIT 2.0-assessed symptom domains in SSc.

We then quantified the magnitude of the association between anti-gephyrin autoantibodies and GI symptom severity. Using univariate logistic regression, we found a significantly higher odds of having anti-gephyrin antibodies among SSc patients with severe constipation [UCLA GIT 2.0; OR 4.79; 95%CI 1.47, 15.61; p=0.009] and severe distention and bloating [OR 3.56; 95%CI 1.12, 11.30; p=0.031] (**Supplemental Table 2**).

### Anti-gephyrin antibody levels associate with the severity of constipation in SSc

We next studied whether anti-gephyrin antibody levels varied across different levels of GI symptom severity (none-to-mild, moderate, and severe), per predefined UCLA GIT 2.0 cutoffs (12). An overall difference between antibody levels across the 3 severity groups of constipation was noted (p=0.013). Furthermore, patients with severe constipation had significantly higher anti-gephyrin antibody levels (β-coeff 0.131, 95%CI 0.04, 0.22; p=0.003; optical density; OD).

To determine whether the association between anti-gephyrin autoantibodies and SSc clinical features would remain after adjusting for potential confounders, multivariable logistic regression models were constructed (**Supplemental Table 2**). Patients with severe constipation (OR 4.74, 95%CI 1.45, 15.47; p=0.010) and severe distention and bloating (OR 3.71, 95%CI 1.16, 11.86; p=0.027) were more likely to be anti-gephyrin antibody positive even after adjustment for disease duration. Furthermore, patients with severe constipation were also significantly more likely to have higher anti-gephyrin antibody levels, even after adjusting for disease duration, though this was not the case for severe distention and bloating (data not shown).

### Anti-gephyrin autoantibodies target myenteric neurons in the gastrointestinal tract

To visualize the cellular targets of anti-gephyrin autoantibodies in the GI tract, anti-gephyrin-positive SSc patient sera (n=5) were used to immunostain ileal LM-MP tissue sections from Wnt1-cre:tdTomato mice. In this system, the red fluorescent protein tdTomato labels all neural crest derivatives, including the ENS. Co-staining performed with a commercial anti-gephyrin antibody confirmed that the patterns observed with patient sera were consistent with gephyrin expression: both labelled enteric neurons in the murine LM-MP, as well as endothelial cells in the murine GI tract (**Figure 2a**). No significant labeling of glial and smooth muscle cells was noted.

**Figure 2:**
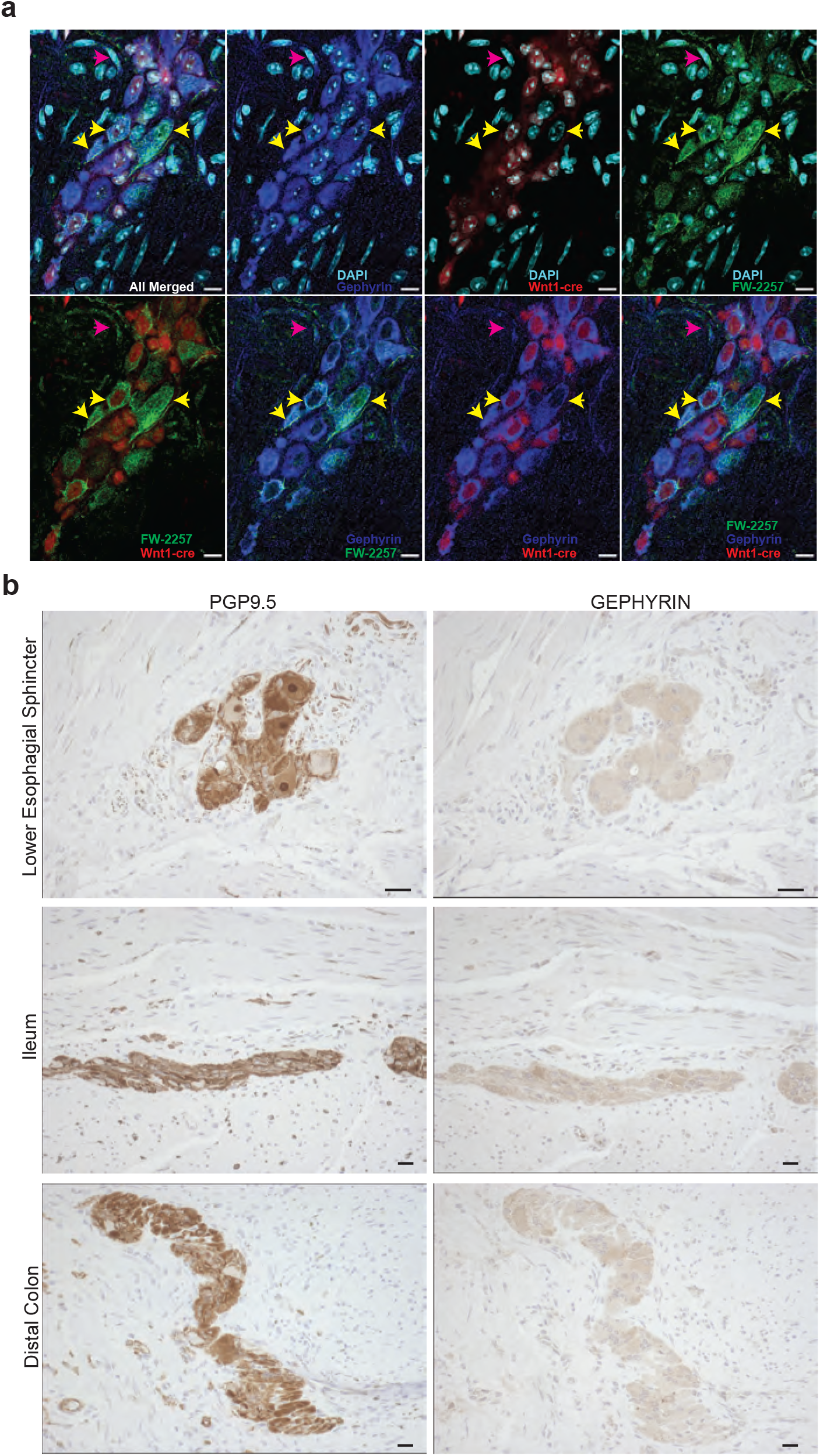
Anti-gephyrin immunoreactivity in murine and human myenteric neurons. (a) Representative image of a murine ileal myenteric ganglion from an adult Wnt1-cre:tdTomato mouse. Merged and other images show Wnt1-derived neural crest-derived cells expressing tdTomato (red) along with immunolabeling with antibodies against gephyrin (blue), patient serum (from patient FW-2257), which contains autoantibodies against gephyrin (green), and nuclear labeling with DAPI (cyan). Yellow arrows denote putative myenteric neuron cells that show co-labeling with the anti-gephyrin commercial antibody and with patient serum. The magenta arrow shows co-staining with the commercial antibody and patient serum in extra-ganglionic cells. The image shown is representative of three 40X fields of views. (b) Representative images of the human myenteric ganglia from tissue sections of various gut regions, namely the lower esophageal sphincter, small intestine (ileum), and distal colon sections. Serial sections were stained with a commercially available antibodies against the pan-neuronal marker PGP9.5 and against gephyrin. These show detectable gephyrin expression in the cells of myenteric ganglia, and appreciable co-localization in serial sections with PGP9.5, a pan-neuronal marker. Images taken at 20X magnification. Scale bar in (a) and (b) = are 10 μm.

To address whether our observation of gephyrin expression by murine enteric neurons extends to human tissue, we performed immunohistochemistry on paraffin sections obtained from distinct human GI tract regions. Gephyrin was detected in PGP9.5 (a pan-neuronal marker) expressing human enteric neurons in various intestinal regions, as well as in GI epithelial cells. Comparable gephyrin expression patterns were noted in tissues from both SSc and normal donors (**Figure 2b**).

## DISCUSSION

Our study is the first to identify gephyrin as a novel target of the autoimmune response in patients with SSc. We determined that the presence and titer of anti-gephyrin antibodies associate strongly with the presence and severity of SSc lower bowel dysfunction. Furthermore, we demonstrate that gephyrin expression is high in the myenteric plexus, which plays a dominant role in regulating lower GI motility.

Anti-gephyrin autoantibodies were first reported in a patient with SSS, a rare disease characterized by dysfunction of the central nervous system (11). Gephyrin is a cytosolic protein that is concentrated at the postsynaptic membrane of inhibitory synapses, where it anchors GABAA and glycine receptors and serves as a critical mediator for neural communications. Given the presenting neurological findings in this SSS patient and the physiological function of gephyrin, the authors proposed that a close link may exist between neurological dysfunction and autoimmunity directed against components of inhibitory neural synapses.

In our study, we determined that anti-gephyrin antibodies are associated with constipation presence and severity in SSc patients. Importantly, receptors for GABA, which require gephyrin for their normal deployment, are expressed by neurons of the myenteric plexus – suggesting that gephyrin expression is possibly functionally important for the normal GABAergic signaling in the myenteric plexus (13). Since patients with severe constipation were significantly more likely to be anti-gephyrin antibody positive, and high levels of anti-gephyrin autoantibodies associated with more severe constipation, it is tantalizing to speculate that autoimmune-mediated disruption in neural communications in the myenteric plexus might lead to impaired bowel transit and prominent constipation (2).

It would be informative to assess whether gephyrin is differentially expressed throughout the human GI tract in a larger SSc tissue sample. Enrichment of gephyrin expression in the myenteric plexus in select gut regions might focus anti-gephyrin immune responses and effector pathways onto the myenteric plexus in these regions. In contrast to the upper gut where the vagal nerve plays a dominant role in controlling GI transit, neural control of the human small and large bowel is predominantly driven by the ENS (2). This distribution of neural control suggests that a disruption of ENS function would have a larger impact on the lower gut compared to the upper gut. Therefore, while gephyrin is expressed in both the upper and lower GI tract, the differential impact of ENS control throughout the gut may help to explain the role in generating the lower bowel dysfunction phenotype in anti-gephyrin antibody positive patients.

Our study has several strengths. Using gut tissue for autoantibody discovery, we are the first to identify anti-gephyrin antibodies in SSc patients. A large, well-characterized cohort of consecutively recruited SSc patients was used to define the prevalence and clinical phenotype associated with this specificity, and to determine that antibody levels associate with disease severity. Gephyrin expression was confirmed in the relevant tissues, notably in the murine and human myenteric neurons of the ENS, which regulate gut motility. Our study also has some limitations. Since it was cross-sectional, further studies are needed to determine whether these antibodies can be utilized to predict phenotype in patients with early disease. While our data provides hypothesis-generating associations suggesting a link with disease mechanism, further studies in animal models are warranted to confirm a causal link.

## Conclusions

These data define novel antibodies in SSc which target gephyrin, a protein that anchors the GABA and glycine receptors at neural synapses and facilitates communications in the ENS. We found that SSc patients with anti-gephyrin antibodies have more severe lower bowel dysfunction, and that antibody levels positively correlate with more severe symptoms. Furthermore, our data demonstrate that gephyrin is expressed in tissues that control lower bowel function. Future research examining the role of anti-gephyrin antibodies as predictors of GI progression, and as mediators of GI progression and dysfunction in SSc are warranted.

## Data Availability

All data produced in the present study are available upon reasonable request to the authors.

## Acknowledgments and affiliations

We thank Michelle Leatherman, Adrianne Woods, Laura Gutierrez, for their contributions to this work.

