## Supplemental Text and Tables for "Anti-Gephyrin antibodies: A novel specificity in patients with systemic sclerosis and lower bowel dysfunction"

**Methods Section**

*Patients*. Disease duration was calculated from the date of the first SSc-associated symptom (Raynaud’s or non-Raynaud’s) to the date of the serum sample. Patients were identified as having limited or diffuse SSc based on the extent of skin involvement (21). The presence of a myopathy was denoted on the basis of: an elevated creatinine phosphokinase with evidence of electromyography (EMG) supportive of myopathy, magnetic resonance imaging (MRI) with evidence of muscle edema, or muscle biopsy consistent with myopathy (18). The muscle severity score was also used to classify the degree of associated proximal muscle weakness and was based on the following scale collected in our database: 0=full strength, 1 = ability to lift upper or lower extremities against gravity with some resistance, 2 = ability to lift upper or lower extremities against gravity only, 3= ability to move upper or lower extremities but not against gravity, and 4= requiring ambulatory aids to walk (22). Cardiac involvement was determined by the Medsger severity scale and was considered present with a score of 1 or greater [0 = normal, 1 = evidence of conduction defect on electrocardiogram or left ventricular ejection fraction (LVEF) of 45-49% on echocardiogram, 2= evidence of arrhythmia on electrocardiogram or LVEF of 40-44%, 3= clinical signs of left or right heart failure or arrhythmia requiring treatment with medication or intervention] (18,22). To capture phenotype, the minimum measurements from the forced vital capacity (FVC) and single breath diffusing capacity of carbon monoxide (DLCO) pulmonary lung function testing and maximum measurements from the estimated right ventricular systolic pressure (RVSP) (measured by transthoracic echocardiogram) were utilized for the analysis (23). Sicca symptoms were defined as the presence of at least one of the following: dry eyes for more than three months, the use of artificial tears three times daily, dry mouth for more than three months, swollen salivary glands, the necessity of liquids for swallowing and/or the sensation of sand in one’s eyes (24).

*Isolation and culture of the longitudinal muscle containing myenteric plexus (LM-MP) from adult murine small intestine*. LM-MP tissue from adult murine ileum was isolated as described, then cultured overnight in defined Neurobasal medium containing B27 growth supplement (14). Typically, each adult murine small intestine yielded 3-to-4 1-cm pieces of ileal tissue. Detergent extracts for use in immunoprecipitations (IPs) for mass spectrometry were made by washing 9 LM-MP peels in phosphate buffered saline (PBS), followed by homogenization in 1 ml of buffer containing 50 mM Tris pH7.4, 150 mM NaCl, 5 mM EDTA, 0.5% Nonidet P40, 0.5% sodium deoxycholate, 0.1% SDS and a protease inhibitor cocktail, followed by centrifugation at 16,000 rpm for 30 min at 4^o^C.

*Identification of antibodies in serum FW-2340.* Detergent extract of LM-MP tissue (0.4 ml, ~560 μg protein) was diluted with 2.6ml Buffer A (1% Nonidet P40, 20 mM Tris pH 7.4, 150 mM NaCl, 1 mM EDTA, and a protease inhibitor cocktail) and precleared with 75 µl of immobilized protein A–agarose (Thermo Scientific). The IP was performed by adding 6 µl of patient serum FW-2340 (see results section for patient details) to the lysate (3 hours, 4^o^C), followed by protein A–agarose. After extensive washing, the IP was further processed at the JHU Proteomics Core facility. Briefly, an on-bead digest was performed with Trypsin/LysC and the resulting peptides were analyzed by reverse phase LC-MS. Eluting peptides were sprayed into a Q-Exactive Plus (QE Plus, Thermo Scientific) mass spectrometer. Isotopically resolved masses were extracted using Proteome Discoverer software and searched using Mascot 2.5.1 through Proteome Discoverer against a mouse protein database. Peptide identifications from Mascot searches were processed within Scaffold (Proteome Software) with display criteria set to 95% confidence for both protein and peptide identifications.

*Assays to validate anti-RUVBL1/2 antibodies.* These were confirmed by IP using radiolabeled full-length ^35^S-methionine–labeled human RUVBL1/2 as input. This was generated from the relevant cDNA (OriGene) by *in vitro* transcription and translation (IVTT), according to the manufacturer’s protocol (Promega). IPs using these products were performed, electrophoresed on 10% sodium dodecyl sulfate (SDS)–polyacrylamide gels and visualized by fluorography as described (15).

*Immunohistochemical staining of human GI tract paraffin sections.* Human autopsy sections were immunostained (overnight, 4^o^C) with an anti-gephyrin rabbit polyclonal antibody (2.7 µg/ml, Proteintech) or an anti-PGP9 rabbit monoclonal antibody (1:250, Abcam) followed by HRP-conjugated anti-rabbit secondary antibody (1:500, Dako). Staining was visualized using the diaminobenzidine substrate chromagen system (Dako). Nuclei were stained using Mayer’s hematoxylin solution (Dako). Light microscopy images were obtained using a Zeiss Axioskop 50 with a Zeiss AxioCam HRC camera and AxioVision 4.9.1 software.

| **Supplemental Table 1. Characteristics of the SSc patients with and without Anti-Gephyrin antibodies in the Johns Hopkins SSc Center all-comers cohort** | | | | |
| --- | --- | --- | --- | --- |
| Clinical and demographic features |  | Anti-Gephyrin antibody positive  (n=16) | Anti-Gephyrin antibody negative  (n=172) | p-value |
| Age at first symptoms, mean (SD) |  | 53 (14) | 58 (13) | 0.20 |
| Disease duration from first SSc-associated symptom (Raynaud’s or non-Raynaud’s) to date of serum sample, median (IQR) |  | 9 (6, 19) | 15 (8, 23) | 0.18 |
| Female sex, % (n) |  | 94 (15) | 86 (148) | 0.70 |
| *Race/Ethnicity* |  |  |  |  |
| White, % (n) |  | 87 (14 ) | 77 (131) | 0.53 |
| Ever smoker, % (n) |  | 25 (4) | 38 (65) | 0.42 |
| *SSc Type* |  |  |  |  |
| Diffuse cutaneous disease, % (n) |  | 63 (10) | 34 (57) | **0.02** |
| Cardiac involvement (≥1), % (n) |  | 25 (2) | 34 (24) | 1.00 |
| Myopathy, % (n) |  | 29 (4) | 22 (34) | 0.52 |
| Sicca symptoms, % (n) |  | 69 (11) | 75 (128) | 0.59 |
| Raynaud’s severity (≥3), % (n) |  | 50 (8) | 60 (103) | 0.44 |
| Lung involvement (≥1), % (n)  Renal crisis, % (n) |  | 20 (2)  0 (0) | 45 (55)  3 (5) | 0.19  1.00 |
| Cancer, % (n) |  | 19 (3) | 19 (33) | 1.00 |
| Telangiectasias, % (n) |  | 88 (14) | 97 (152) | 0.10 |
| Tendon friction rub, % (n) |  | 38 (6) | 19 (32) | 0.07 |
| Calcinosis, % (n) |  | 38 (6) | 45 (78) | 0.61 |
| Dead, % (n) |  | 0 (0) | 4 (6) | 1.00 |
| *Pulmonary function parameters* |  |  |  |  |
| FVC, % (n) |  | 86 (19) | 73 (19) | **0.01** |
| DLCO, % (n) |  | 80 (15) | 64 (20) | **<0.01** |
| RVSP by echo (mmHg), mean (SD) |  | 34 (4) | 37 (12) | 0.68 |
| *Antibodies, % (n)* |  |  |  |  |
| Scl-70 (i.e. Topoisomerase-1) |  | 19 (3) | 19 (32) | 1.00 |
| Centromere (CENP) |  | 31 (5) | 30 (51) | 1.00 |
| RNA polymerase-3 |  | 25 (4) | 14 (23) | 0.21 |
| U3RNP |  | 0 (0) | 4 (7) | 1.00 |
| PMScl |  | 13 (2) | 7 (12) | 0.34 |
| ThTo |  | 0 (0) | 2 (4) | 1.00 |
| Ku |  | 6 (1) | 3 (5) | 0.42 |
| *SD = standard deviation; IQR = interquartile range; FVC = Forced Vital Capacity & DLCO = Diffusion Capacity on PFT* | | | | |

| **Supplemental Table 2. Statistical models evaluating the association between anti-Gephyrin antibody positive systemic sclerosis patients and clinical features** | | | | |
| --- | --- | --- | --- | --- |
|  | Unadjusted Logistic Regression | | Adjusted Logistic Regression* | |
| *Characteristic* | *OR* | *95% Confidence Interval* | *OR* | *95% Confidence Interval* |
| Diffuse cutaneous disease | 3.27 | 1.13-9.46 | 2.85 | 0.92-8.79 |
| FVC (% predicted) | 1.04 | 1.01-1.08 | 1.04 | 1.01, 1.08 |
| DLCO (% predicted) | 1.04 | 1.01-1.07 | 1.04 | 1.01, 1.07 |
| Constipation, severe** | 4.79 | 1.47-15.61 | 4.74 | 1.45, 15.47 |
| Distention/ bloating, severe** | 3.56 | 1.12-11.30 | 3.71 | 1.16, 11.86 |
| *FVC = Forced Vital Capacity on PFT; DLCO = Diffusion Capacity on PFT; *adjusted for disease duration **UCLA GIT 2.0* | | | | |
